# An inactivated SARS-CoV-2 vaccine is safe and induces humoral and cellular immunity against virus variants in healthy children and adolescents in Chile

**DOI:** 10.1101/2022.02.15.22270973

**Authors:** Jorge A Soto, Felipe Melo-González, Cristián Gutierrez-Vera, Bárbara M Schultz, Roslye V Berríos-Rojas, Daniela Rivera-Pérez, Alejandro Piña-Iturbe, Guillermo Hoppe-Elsholz, Luisa F Duarte, Yaneisi Vázquez, Daniela Moreno-Tapia, Mariana Ríos, Pablo A Palacios, Richard Garcia-Betancourt, Álvaro Santibañez, Constanza Mendez, Benjamín Diethelm-Varela, Patricio Astudillo, Mario Calvo, Antonio Cárdenas, Marcela González, Macarena Goldsack, Valentina Gutiérrez, Marcela Potin, Andrea Schilling, Lorena I Tapia, Loreto Twele, Rodolfo Villena, Alba Grifoni, Alessandro Sette, Daniela Weiskopf, Rodrigo A Fasce, Jorge Fernández, Judith Mora, Eugenio Ramírez, Aracelly Gaete-Argel, Mónica Acevedo, Fernando Valiente-Echeverría, Ricardo Soto-Rifo, Angello Retamal-Díaz, Nathalia Muñoz-Jofré, PedCoronaVac03CL Study Group, Xing Meng, Qianqian Xin, Eduardo Alarcón-Bustamante, José V González-Aramundiz, Nicole Le Corre, María Javiera Álvarez, Pablo A González, Katia Abarca, Cecilia Perret, Leandro J Carreño, Alexis M Kalergis, Susan M Bueno

**Affiliations:** Millennium Institute on Immunology and Immunotherapy, Santiago, Chile; Departamento de Genética Molecular y Microbiología, Facultad de Ciencias Biológicas, Pontificia Universidad Católica de Chile, Santiago, Chile; Programa de Inmunología, Instituto de Ciencias Biomédicas, Facultad de Medicina, Universidad de Chile, Santiago, Chile; Departamento de Enfermedades Infecciosas e Inmunología Pediátrica, División de Pediatría, Escuela de Medicina, Pontificia Universidad Católica de Chile, Santiago, Chile; Universidad Austral de Chile, Valdivia, Los Ríos, Chile; Departamento de Ciencias Médicas, Facultad de Medicina y Odontología, Universidad de Antofagasta, II Región, Chile; Hospital Dr. Gustavo Fricke, V Región, Chile; Departamento de Pediatría, Universidad de Valparaíso, V Región, Chile; Unidad de Infectología Pediátrica, Servicio de Pediatría, Complejo Asistencial Dr. Sótero del Río, Santiago, Chile; Clínica San Carlos de Apoquindo, Red de Salud UC Christus, Santiago, Chile; Facultad de Medicina, Clínica Alemana, Universidad del Desarrollo, Santiago, Chile; Departamento de Pediatría y Cirugía Infantil Norte, Hospital Roberto del Río; Programa de Virología, Instituto de Ciencias Biomédicas, Facultad de Medicina, Universidad de Chile, Santiago, Chile; Hospital Puerto Montt, X Región, Chile; Hospital Exequiel González Cortés, Santiago, Chile; Center for Infectious Disease and Vaccine Research, La Jolla Institute for Immunology (LJI), La Jolla, CA 92037, USA; Department of Medicine, Division of Infectious Diseases and Global Public Health, University of California, San Diego (UCSD), La Jolla, CA 92037, USA; Departamento de Laboratorio Biomédico, Instituto de Salud Pública de Chile, Santiago, Chile; Laboratorio de Virología Molecular y Celular, Programa de Virología, Instituto de Ciencias Biomédicas, Facultad de Medicina, Universidad de Chile, Santiago, Chile; Departamento de Biotecnología, Facultad de Ciencias del Mar y de Recursos Biológicos, Universidad de Antofagasta, II Región, Chile; Departamento de Tecnología Médica, Facultad de Ciencias de la Salud, Universidad de Antofagasta, II Región, Chile; SINOVAC Biotech, Beijing, China; Faculty of Mathematics, Department of Statistics, Pontificia Universidad Católica de Chile, Santiago, Chile; Millennium Nucleus on Intergenerational Mobility: From Modelling to Policy, MOVI, Santiago, Chile; Interdisciplinary Laboratory of Social Statistics, Santiago, Chile; Departamento de Farmacia, Facultad de Química y de Farmacia, Pontificia Universidad Católica de Chile, Santiago, Chile; Departamento de Endocrinología, Facultad de Medicina, Escuela de Medicina, Pontificia Universidad Católica de Chile, Santiago, Chile

**Keywords:** CoronaVac^®^, Phase 3 clinical trial, Pediatric, SARS-CoV-2, COVID-19, Vaccines, Variants of Concern

## Abstract

**Background:** Multiple vaccines against SARS-CoV-2 have been evaluated in clinical trials, but very few include the pediatric population. The inactivated vaccine CoronaVac^®^ has shown to be safe and immunogenic in a phase 1/2 clinical trial in a pediatric cohort in China. This study is an interim safety and immunogenicity report of a phase 3 clinical trial for CoronaVac^®^ in healthy children and adolescents in Chile.

**Methods:** Participants aged 3 to 17 years old received two doses of CoronaVac^®^ in a four-week interval. Local and systemic adverse reactions were registered in 699 participants that received the first dose and 381 that received the second dose until December 31^st^, 2021. Whole blood samples were collected from 148 participants for humoral and cellular immunity analyses.

**Results:** The primary adverse reaction reported after the first and second dose was pain at the injection site. The adverse reactions observed were primarily mild and local, and no severe adverse events were reported. Four weeks after the second dose, a significant increase in the levels of total and neutralizing antibodies was observed. Increased activation of specific CD4^+^ T cells was also observed four weeks after the second dose. Although antibodies induced by vaccination neutralize variants Delta and Omicron, titers were lower than the D614G variant. Importantly, comparable T cell responses were detected against these variants of concern.

**Conclusions:** CoronaVac^®^ is safe and immunogenic in subjects aged 3-17 years old and is thus likely to confer protection against infection caused by SARS-CoV-2 variants in this target population.

## Introduction

Severe acute respiratory syndrome coronavirus-2 (SARS-CoV-2), the etiological agent of coronavirus disease 2019 (COVID-19), has been responsible for over 307.2 million cases and 5.4 millions deaths worldwide (at February 2^nd^, 2022)^1^. Currently, multiple vaccines based on different platforms have been developed to reduce the transmission and severity of COVID-19 ^2^. Clinical trials for these vaccines have been conducted in different countries in healthy adults, but relatively few have been performed or reported in adolescents and children ^3–5^. Although adolescents and children are usually asymptomatic upon infection with SARS-CoV-2 and mostly develop mild disease, they still can be hospitalized needing intensive care and even mechanical ventilation ^6^. In addition, in rare cases, they can suffer a disease called multisystem inflammatory syndrome in children (MIS-C)^7^. Thus, more studies on SARS-CoV-2 vaccines are needed in children and adolescents to understand better the immune responses associated with vaccination.

The inactivated SARS-CoV-2 vaccine CoronaVac^®^, developed by Sinovac Life Sciences Co., Ltd. (Beijing, China), has been approved by the World Health Organization (WHO) for its use in adults against COVID-19 based on several clinical trials that have proven its safety, immunogenicity, and efficacy ^8–12^. This vaccine was tested in clinical trials in adults in several countries, including China, Brazil, Turkey, and Chile. Clinical trials in these countries have shown that CoronaVac^®^ promotes anti-Spike IgG antibodies and anti-Spike Receptor-Binding Domain (RBD) neutralizing antibodies, together with cellular immune responses against SARS-CoV-2 antigens in healthy adult participants ^13^. A clinical trial conducted in China with CoronaVac^®^ also showed favorable safety and immunogenicity results in children and adolescents aged between 3-17 years old, which displayed neutralizing antibodies titers against SARS-CoV-2 after immunization ^3^. Similarly, other vaccines such as Pfizer BNT162b2 and Moderna mRNA-1273 have been tested in children between 6-11 years old and adolescents, showing to be safe and to induce neutralizing antibodies against SARS-CoV-2 ^4,5^. However, these reports lack a characterization of the cellular immune responses elicited in vaccinated children and adolescents after immunization, as well as the characterization of the neutralizing capacity of antibodies against SARS-CoV-2 variants of concern. Here, we further characterize the immune responses elicited in participants aged between 3 and 17 years old four weeks after the second dose of CoronaVac^®^ applied in a 4 week interval (or 0-28-day vaccination schedule), demonstrating that this vaccine is safe and elicit significant levels of both humoral and cellular immunity in adolescents and children.

## Materials and methods

### Study design

This study is a global multi-center, randomized, double-blinded, and placebo-controlled phase 3 clinical trial that aims to assess the safety, efficacy, and immunogenicity of CoronaVac^®^ among children aged six months to 17 years. Four countries participated in this study, including South Africa, Malaysia, Philippines, and Chile (clinicaltrials.gov #NCT04992260). This report will only focus on the study performed in Chile for participants that received CoronaVac®. In Chile, this trial has been conducted at eleven different sites, eight in the center of the country (seven in Santiago and one in Valparaiso), two in the South (Puerto Montt and Valdivia), and one in the North (Antofagasta) of Chile. The study protocol was conducted according to the current Tripartite Guidelines for Good Clinical Practices, the Declaration of Helsinki ^14^, and local regulations. This trial was approved by each Institutional Ethical Committee (ID 210616012) and the Chilean Public Health Institute (ISP Chile, number N° 20674/21). Written informed consent was obtained from the parent(s) or legal representative(s) of the child before enrollment. An assent was obtained in individuals from 7 years old. Participants did not receive any payment for their participation. The study included 3-17 years old children and adolescents, who were inoculated with two doses of 3µg (600SU) of CoronaVac^®^ in a 4-week interval (0-28 schedule) (**Figure 1A**). Exclusion criteria included, among others, history of confirmed symptomatic SARS-CoV-2 infection, pregnancy, allergy to vaccine components, and immunocompromised condition. Well controlled medical conditions were allowed. A complete list of inclusion/exclusion criteria is provided in supplementary materials.

**Figure 1.**
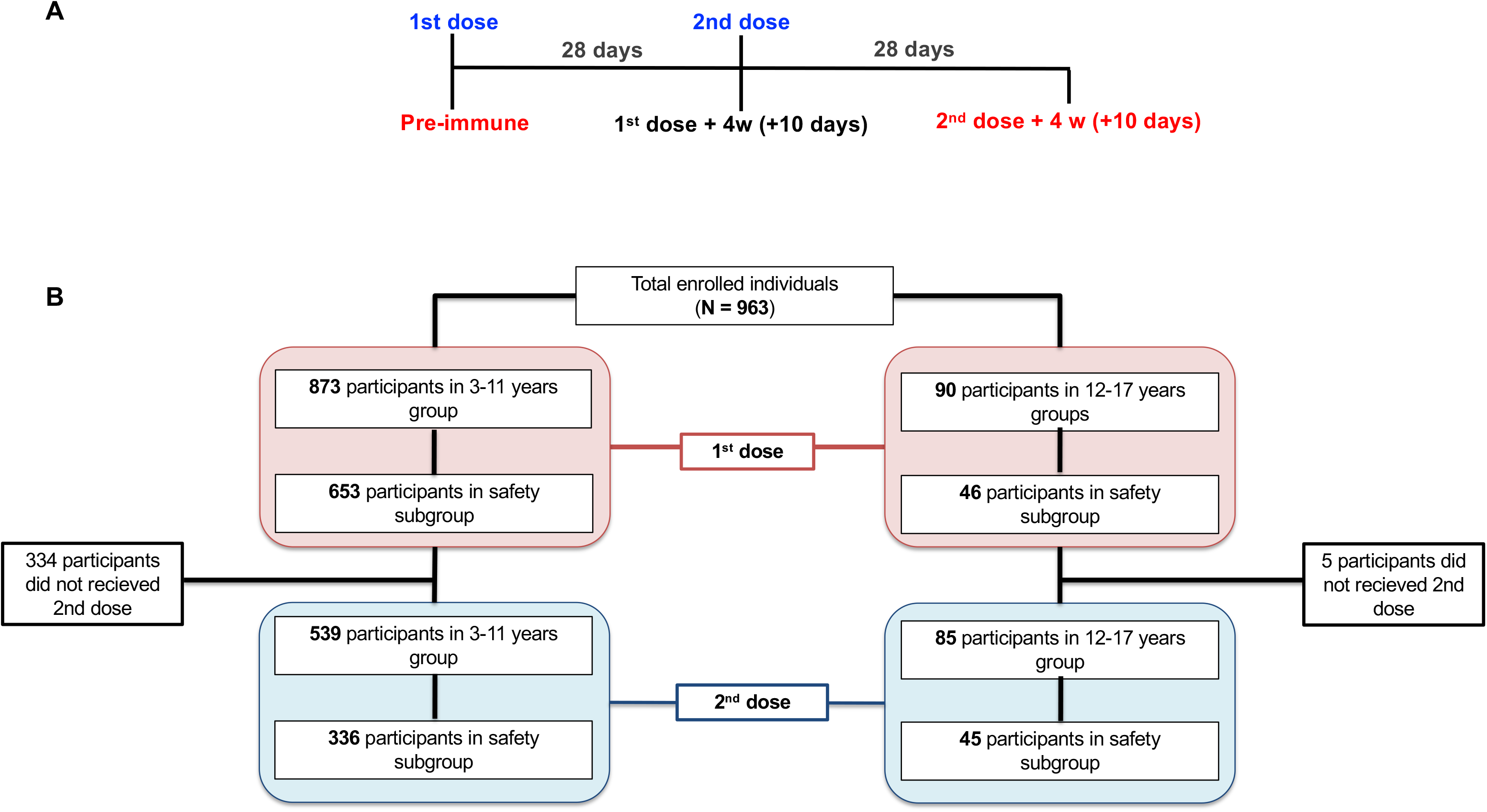
Study profile, enrolled participants and cohort included in this study from September 10^th^ to December 31^st^, 2021. **A.** Timeline of the vaccination schedule and sample collection. Text in red denotes timepoints at which blood draws occurred. **B.** Study profile of subjects that received 1 dose (orange boxes) and 2 doses (light blue boxes) until December 31^st^, 2021, by age and safety group.

### Study population and outcomes

Participants were assigned to the 3-5 (children), 6-11 (children), and 12-17 (adolescents) age group and immunogenicity subgroup, safety subgroup, or non-subgroup. For the present study, we combined the 3-11 age group (children) (**Figure 1B**). Safety group includes registration of every local and systemic non-immediate adverse events (AE) in the seven days after vaccination and any other AE until 28 days after each dose. For all participants, immediate AE (30 min post-vaccination) and serious adverse events (SAE), and adverse events of special interest (AESI) were recorded.

The study aims were to evaluate the immunogenicity of CoronaVac^®^ in a subgroup of participants 4 weeks after 2 doses and the frequency of solicited immediate (first 30 min post-dose) and non-immediate adverse events (AEs) that occur during seven days after each dose, stratified by age group (3-11 and 12-17 years old), and the frequency of SAE/AESI and any other AE occurring 28 days after each dose, and the frequency of any SAE/AESI occurring 12 months after the second dose.

### Sample collection

Subjects enrolled in one specific clinical center (CL01, Marcoleta) were assigned to the immunogenicity branch. Blood samples were obtained in heparinized tubes before administration of the first dose (pre-immune) and four weeks after the second dose, as described in **Suppl. Figure 1**. Samples were used to obtain plasma and peripheral blood mononuclear cells (PBMCs) and stored at - 80°C (plasma) and -170°C (PBMCs) until humoral and cellular immunity analyses were performed. The sample size included in each experimental analyses is described in **Suppl. Figure 1**.

### Evaluation of specific antibody levels and their neutralization capacity

IgG anti-S1-RBD of SARS-CoV-2 were tested using ADVIA Centaur® XP SARS-CoV-2 IgG (sCOVG, Siemens)^15,16^, an automated two-step sandwich antibody-binding immunoassays using indirect chemiluminescence. sCOVG was used for quantitative detection expressed in BAU/mL after interpolating the WHO standard NIBSC code 20/136 calibration 3.

The presence of circulating antibodies able to block the interaction of the RBD of the S1 subunit of the SARS-CoV-2 Spike protein with the recombinant human Angiotensin 2 Receptor (hACE2) was evaluated using a surrogate virus neutralization test (sVNT) (Genscript Cat#L00847-A). Two-fold serial dilutions were prepared for each sample, starting at a 4-fold until reaching a 512-fold dilution, and the assay was performed according to manufacturers’ instructions ^17^. The end titer of neutralizing antibodies was assigned as the last fold dilution that displayed a cut-off over 30% of inhibition. Seropositivity designation and transformation to WHO Arbitrary Units was described previously^25^. Samples with undetermined concentration at the lowest dilution tested (1:4) were assigned the lower limit of quantification (16.4 IU).

Conventional virus neutralization tests (cVNT) were performed as previously reported ^17^. Briefly, Vero E6 cells were infected with a SARS-CoV-2 strain obtained by viral isolation in tissue cultures (33782CL-SARS-CoV-2 strain, D614G variant). Neutralization assays were carried out by the reduction of cytopathic effect (CPE) in Vero E6 cells (ATCC CRL-1586). The titer of neutralizing antibodies was defined as the highest plasma dilution that neutralized virus infection, at which the CPE was absent as compared with the virus control wells (cells with CPE). Vero E6 cells were seeded in 96-well plates (4×10^4^ cells/well). For neutralization assays, 100 µL of 33782CL-SARS-CoV-2 (at a dose of 100 TCID_50_) were incubated with serial dilutions of heat-inactivated sera samples from participants (dilutions of 1:4, 1:8, 1:16, 1:32, 1:64, 1:128, 1:256, and 1:512) from participants for 1h at 37 °C. Cytopathic effect on Vero E6 cells was analyzed seven days after infection.

A pseudotyped virus neutralization test (pVNT) assay was performed to assess the neutralization capacity of the antibodies against SARS-CoV-2 variants of concern (VOC). As previously reported ^19^, an HIV-1 backbone expressing firefly luciferase as a reporter gene and pseudotyped with the SARS-CoV-2 spike glycoproteins (HIV-1-SΔ19) from lineage B.1 (D614G) or variants Delta (T19R, del157/158, L452R, T478K, D614G, P681R, D950N) and Omicron (A67V, ΔH69-V70, T95I, Y145D, ΔG142 -V143-Y144, ΔN211, EPE 213-214, G339D, S371L, S373P, S375F, K417N, N440K, G446S, S477N, T478K, E484A, Q493R, G496S, Q498R, N501Y, T547K, D614G, H655Y, N679K, P681H, N764K, N865K, Q954H, N969K, L981F) was prepared as previously described ^20^. Plasma samples were two-fold diluted, starting at 1:10 or 1:4, and the estimation of the ID80 was obtained using a 4-parameter nonlinear regression curve fit measured as the percent of neutralization determined by the difference in average relative light units (RLU) between test samples and pseudotyped virus controls. Data analyses and statistical analyses were carried out using GraphPad Prism v9.

### Determination of SARS-CoV-2 specific T cell responses

In order to assess the cellular immune response in vaccinated children and adolescents, PBMCs of sixty participants were stimulated with six Mega Pools (MPs) of peptides derived from the proteome of SARS-CoV-2, including peptides from the S protein of SARS-CoV-2 (MP-S) ^21^, the remaining proteins of the viral particle (excluding S protein peptides) (MP-R) ^21^, peptides from the M protein (Miltenyi, Cat#130-126-702), peptides from the N protein (Miltenyi, Cat#130-126-698) and MHC-I restricted peptides from the whole proteome of SARS-CoV-2 (MP-CD8-A and MP-CD8-B) ^21^. MP of peptides from the S protein of SARS-CoV-2 VOC Delta and Omicron were provided by La Jolla Institute for Immunology ^22^. Positive and negative controls were included in each assay. The number of Spot Forming Cells (SFC) for IFN-γ and IL-4 were determined by ELISPOT, and the expression of Activation-Induced Markers (AIM^+^) and memory markers by T cells was evaluated by flow cytometry using a LSR Fortessa X-20 flow cytometer (BD Biosciences). Assays were performed according to the manufacturer’s instructions and as reported previously^17^.

Supernatants from PBMCs stimulated with SARS-CoV-2 MPs for 20h were evaluated using the Luminex^®^ technology (R&D systems, USA) to assess IL-2 and IFN-γ production. Briefly, supernatants of samples stored at -80°C were thawed at room temperature and diluted 1:2 before analysis. After 2 h incubation with spectrally encoded beads, coated with analyte-specific biotinylated primary antibodies, the samples were incubated with streptavidin R-phycoerythrin and analyzed using a Luminex 200 xMap multiplex system (Luminex Corporation, Austin, TX). According to the manufacturer’s instruction the detection limit for the cytokines measured ranged from 4.2 to 13,390 pg/mL.

### Statistical analyses

Statistical differences for the immunogenicity results were assessed using the use of Wilcoxon test analyzed data to compare the levels of antibodies four weeks after the second dose against the pre-immune levels, whereas the Mann-Whitney test was used to compare the level of antibodies four weeks after the second dose between both age groups (Total IgG, sVNT, pVNT, and cVNT). A two-way ANOVA was used for cellular immune response to compare the percentage of AIM^+^, memory AIM^+^ CD4^+^ T cells, and cytokines secretion four weeks after the second dose against the pre-immune levels in both age groups. The significance level was set at 0.05 for all the analyses. All data were analyzed with GraphPad Prism 9.0.1.

## Results

### Population included in the study

Nine hundred sixty-three participants were recruited between September 10th and December, 31th, 2021, 482 of them are male (51.1%), average age 6.35 years old (SD 3.12). **Figure 1B** shows the enrolled population and distribution by age, dose and safety group.

### Safety, and adverse events identified in children and adolescents vaccinated with CoronaVac^®^

#### Immediate adverse events

In the 30 min post-vaccination, local pain was reported by 3.8% and 1.7% of participants 3-11 years old after the first and second dose, respectively, and in 2.2% and 8.2% of adolescents. Pain was statistically significantly higher in adolescents than in children after the second dose (p= 0.002791). The rest of local AEs were reported in 2% or fewer participants, without age or dose differences (**Suppl. Table 1**).

Systemic immediate AE were reported in less than 1% of 3-11 years old participants. Meanwhile, adolescents reported 2.2% and 1.2% headaches after the first and second dose, respectively. They reported no other systemic AEs after the first dose, and one adolescent reported auto-limited pruritus after the second dose (**Suppl. Table 2**).

#### Non-immediate adverse events

Only the safety group reported non-immediate AEs (**Figure 1B**). The most frequent local non-immediate AEs was pain, observed in around 15% of 3-11 years old children after the first dose and in 8% after the second dose (p=0.003). In adolescents, pain was reported in around 25% after each dose, being significantly higher than children after the second dose (p=0.0063). The rest of the local AEs were reported in less than 5% of 3-11 years old and in less than 10% of adolescents. Most local AEs resolved in 2 days (**Table 1**).

**Table 1.**
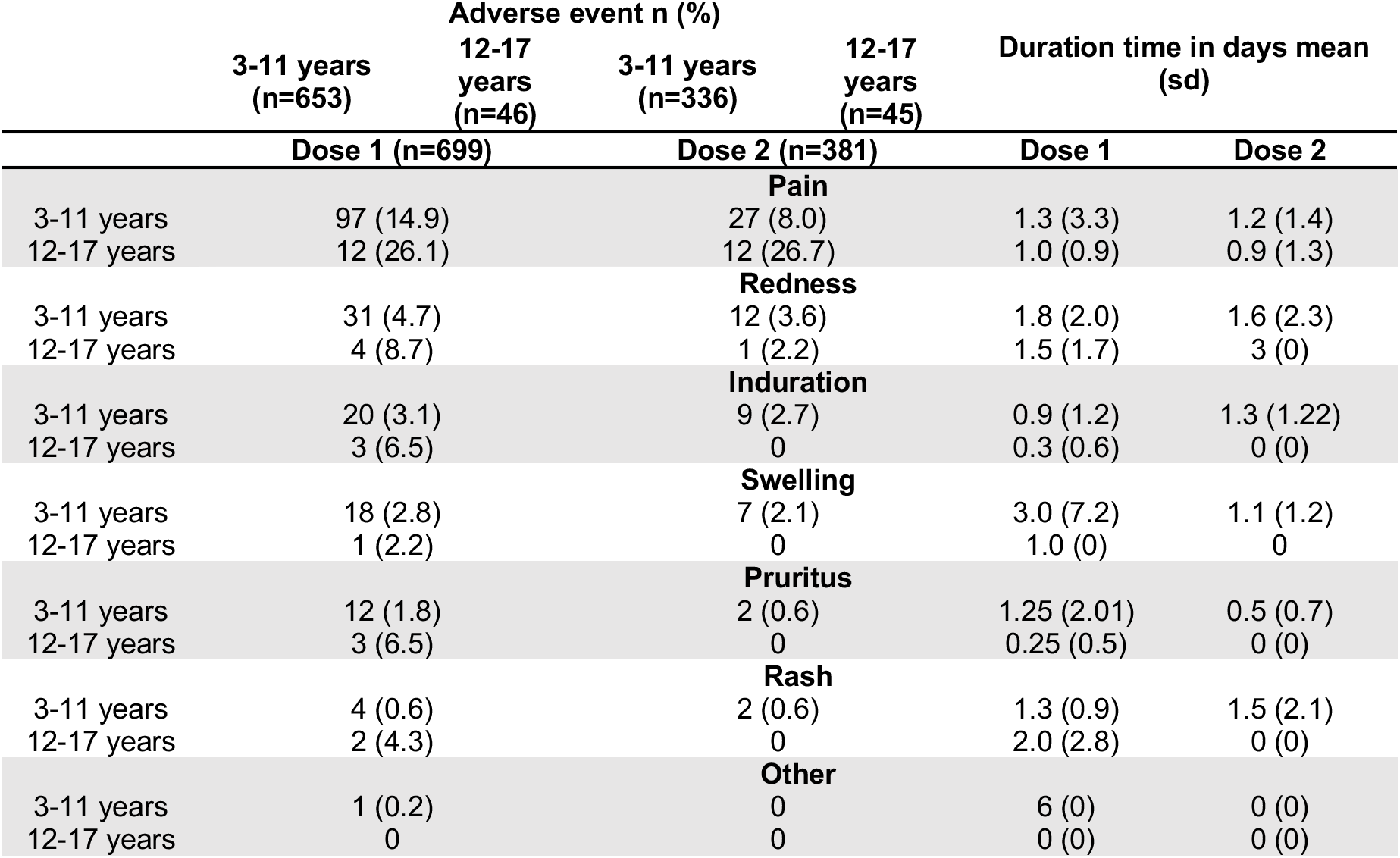
Frequency and duration of local non-immediate adverse events by dose and age group.

Systemic AEs were reported at a frequency lower than 10% each. Headache was the most common AE in adolescents; meanwhile, it was fever in 3-11 years old children. In these age groups, fever was reported in 9 and 7% after the first and second dose, respectively, but in just one adolescent after the first dose. The rest of the systemic AEs were reported in less than 10% of the vaccinated subjects (**Table 2**). Comparison by age and dose showed significantly higher headache in adolescents than in younger children after both doses (p=0.00016 and 0.0028), and higher fatigue in children 3-11 years old after the first than after the second dose (p=0.0117). The severity of systemic AE was grade 1 in 62-79% of participants and grade 3 in only 1.7-2.7%. There was no grade 4 AE (**data not shown**).

**Table 2.** Frequency and duration of systemic adverse events by dose and age group.

| Adverse event n (%) |  |  |  |  | Duration time in days<br>mean (sd) |  |
| --- | --- | --- | --- | --- | --- | --- |
|  | 3-11 years<br>(n=653) | 12-17 years<br>(n=46) | 3-11 years<br>(n=336) | 12-17 years<br>(n=45) |  |  |
|  | Dose 1 (n=699) |  | Dose 2 (n=381) |  | Dose 1 | Dose 2 |
| Fever |  |  |  |  |  |  |
| 3-11 years | 57 (8.7) |  | 22 (6.5) |  | 1.1 (4.0) | 1.6 (2.2) |
| 12-17 years | 1 (2.2) |  | 0 |  | 1 (0) | 0 (0) |
| Fatigue |  |  |  |  |  |  |
| 3-11 years | 41 (6.3) |  | 8 (2.4) |  | 2.1 (5.2) | 1.6 (1.4) |
| 12-17 years | 5 (10.9) |  | 6 (13.3) |  | 0.2 (0.4) | 1.7 (1.6) |
| Headache |  |  |  |  |  |  |
| 3-11 years | 39 (6.0) |  | 13 (3.9) |  | 1.1 (2.18) | 1.8 (1.8) |
| 12-17 years | 11 (23.9) |  | 4 (8.9) |  | 0.8 (1.2) | 0.5 (1) |
| Diarrhea |  |  |  |  |  |  |
| 3-11 years | 33 (5.1) |  | 10 (3.0) |  | 1.2 (1.5) | 1.7 (1.7) |
| 12-17 years | 2 (4.3) |  | 1 (2.2) |  | 2.5 (3.5) | 2 (0) |
| Vomiting |  |  |  |  |  |  |
| 3-11 years | 26 (4.0) |  | 13 (3.9) |  | 1.1 (2.5) | 0.6 (1.2) |
| 12-17 years | 0 |  | 0 |  | 0 (0) | 0 (0) |
| Muscle pain |  |  |  |  |  |  |
| 3-11 years | 18 (2.8) |  | 8 (2.4) |  | 1.4 (1.9) | 0.3 (0.5) |
| 12-17 years | 4 (8.7) |  | 3 (6.7) |  | 2.3 (2.6) | 2 (1) |
| Skin or mucosa abnormality |  |  |  |  |  |  |
| 3-11 years | 17 (2.6) |  | 2 (0.6) |  | 4.3 6.7) | 5 (3.7) |
| 12-17 years | 3 (6.5) |  | 0 |  | 3 (0) | 0 (0) |
| Anorexia |  |  |  |  |  |  |
| 3-11 years | 16 (2.5) |  | 8 (2.4) |  | 1.8 (2.1) | 0.7 (0.9) |
| 12-17 years | 1 (2.2) |  | 1 (2.2) |  | 0 (0) | 2 (0) |
| Allergic reaction |  |  |  |  |  |  |
| 3-11 years | 7 (1.1) |  | 1 (0.3) |  | 3.5 (2.4) | - |
| 12-17 years | 0 |  | 0 |  | 0 (0) | 0 |
| Nausea |  |  |  |  |  |  |
| 3-11 years | 6 (0.9) |  | 2 (0.6) |  | 0.5 (0.8) | 0 (0) |
| 12-17 years | 2 (4.3) |  | 0 |  | 10.7<br>(17.6) | 0 (0) |
| Other |  |  |  |  |  |  |
| 3-11 years | 133 (20.4) |  | 51 (15.2) |  | 3.8 (4.0) | 4.3 (3.7) |
| 12-17 years | 10 (21.7) |  | 12 (26.7) |  | 2.3 (3.3) | 3.3 (3.3) |

There was just one no related SAE reported in the period (a 3-year-old participant hospitalized for 24 hours due to influenza A infection).

### Two doses of CoronaVac^®^ induce a significant increase in antibody titers with neutralizing capacity in children and adolescents

Ninethy-two participants from the immunogenicity branch, who received two doses of the CoronaVac^®^, were included in this study (**Suppl. Figure 1**). Samples analyzed were obtained before vaccination (pre-inmmune) and four weeks after the second dose. We evaluated the induction of IgG against RBD-S1 of SARS-CoV-2 by chemoelectroluminescence (**Figure 2A-C**), which are significantly increased following two doses of CoronaVac as compared to the pre-immune sample. Accordingly, we detected significant neutralizing capacity in plasma obtained from the 3-11 age group (GMU 713.1, 95% CI=565.8-898.8) and the 12-17 years old group (GMU 492.2, 95% CI=342.0-708.3) following two doses of CoronaVac^®^ four weeks after the second dose, when expressed as international units of WHO (**Figure 2D-F**), which is in line with previous reports in adult cohorts^13^. In addition, the seropositivity reached 100% for the samples analyzed four weeks after the second dose in both groups. Similarly, when analyzing neutralization against the live virus using a cVNT, we observed a significant increase in both age groups (**Suppl. Figure 2A and B)**: GMT 128, 95% CI= 74.8-219.2 for age group 3-11 years old and GMT 34.02, 95% CI= 18.1-64.0 for age group 12-17 years old (**Table 3**). A significant difference in the titers of neutralizing antibodies is observed between both age groups (**Figure 2C** and **Suppl. Figure 2C).**

**Figure 2.**
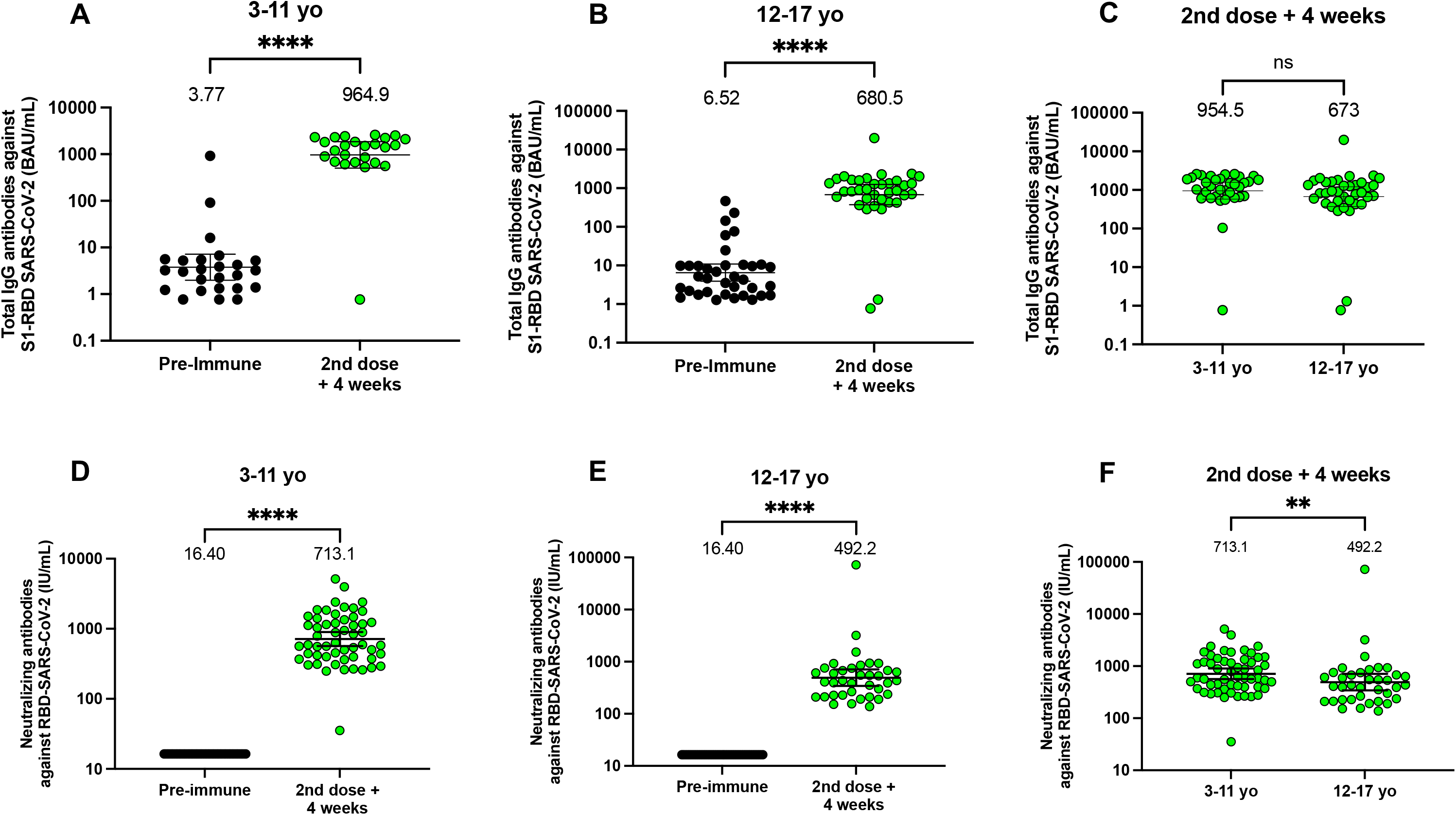
CoronaVac^®^ immunization induces anti-S1-RBD antibodies with neutralizing capacities in children and adolescents after two vaccine doses. **A-C.** Total IgG anti-S1-RBD antibodies were detected by chemoelectroluminiscence in plasma of participants immunized with CoronaVac^®^. Results were obtained from (**A**), twenty-five participants aged 3-11 years old and (**B**) thirty-six participants aged 12-17 years old. (**C**) Comparison of total IgG andti-S1-RBD in both age groups, four weeks after the second dose. **D-F** Neutralizing antibodies were detected in plasma of participants immunized with CoronaVac^®^ using a surrogate Viral Neutralization Test (sVNT), which quantifies the interaction between S1-RBD and hACE2 on ELISA plates. Results were obtained from (**D**) fifty-five participants aged 3-11 years old and (**E**) thirty-six participants aged 12-17 years old. (**F**) Both age groups four weeks after the second dose were compared. Data is represented as WHO arbitrary units/mL, the numbers above each set of individual data points show the Geometric Mean Units (GMU) and the error bars indicate the 95% CI. A Wilcoxon test analyzed data to compare the levels of antibodies four weeks after the second dose against the pre-immune whereas a Mann-Whitney test was used to compare the level of antibodies four weeks after the second dose in both age groups. **p<0.005, ****p<0.0001, n.s. non-significant.

**Table 3:**
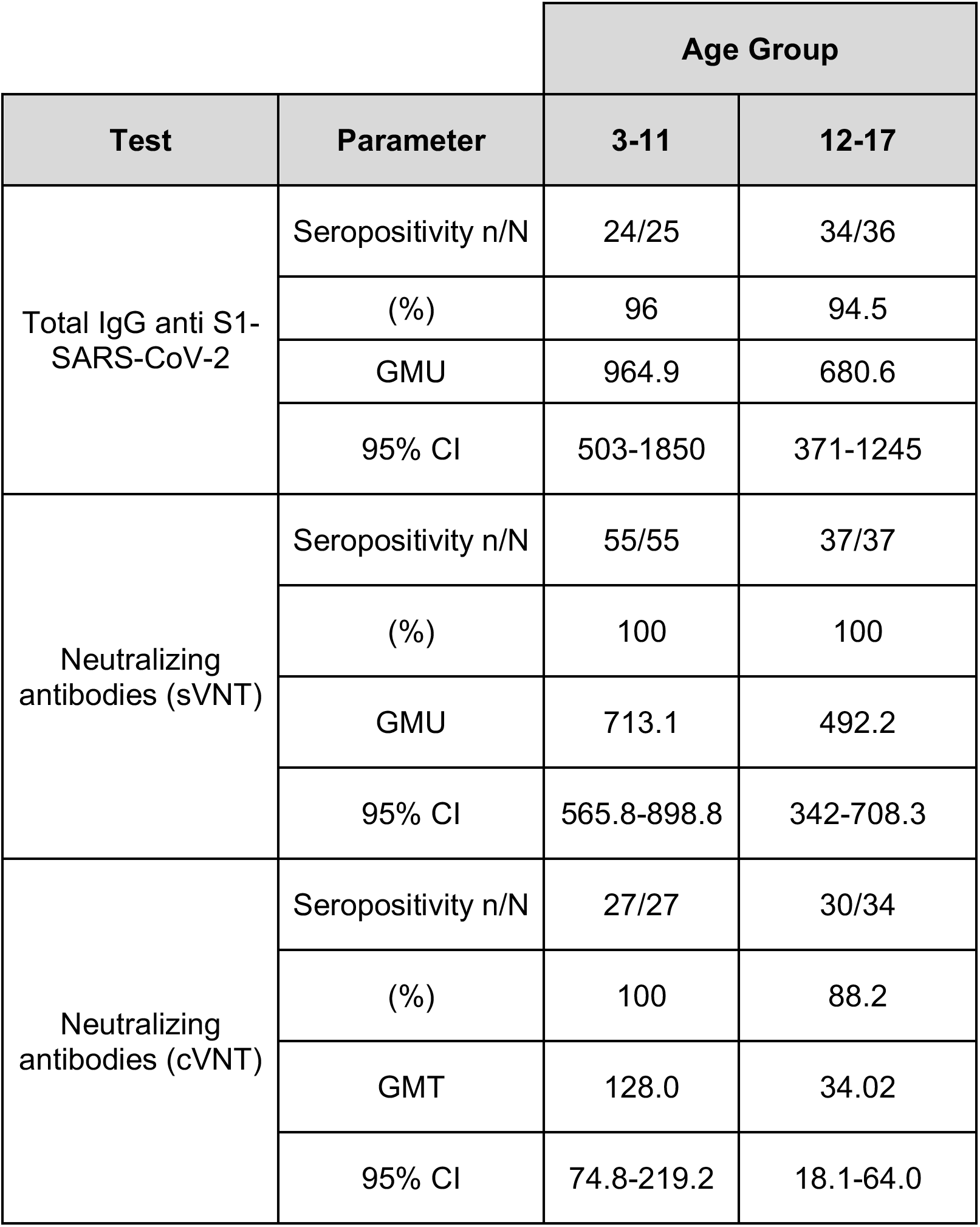
Seropositivity rates, Geometric Mean Units (GMU) or titers (GMT) of circulating antibodies against SARS-CoV-2.

|  |  | Age Group |  |
| --- | --- | --- | --- |
| Test | Parameter | 3-11 | 12-17 |
| Total IgG anti S1-SARS-CoV-2 | Seropositivity n/N | 24/25 | 34/36 |
|  | (%) | 96 | 94.5 |
|  | GMU | 964.9 | 680.6 |
|  | 95% CI | 503-1850 | 371-1245 |
| Neutralizing antibodies (sVNT) | Seropositivity n/N | 55/55 | 37/37 |
|  | (%) | 100 | 100 |
|  | GMU | 713.1 | 492.2 |
|  | 95% CI | 565.8-898.8 | 342-708.3 |
| Neutralizing antibodies (cVNT) | Seropositivity n/N | 27/27 | 30/34 |
|  | (%) | 100 | 88.2 |
|  | GMT | 128.0 | 34.02 |
|  | 95% CI | 74.8-219.2 | 18.1-64.0 |

### Two doses of CoronaVac^®^ induce a robust activation and memory population of CD4^+^ T cells in children and adolescents

We also analyzed the cellular immune responses following two doses of CoronaVac^®^ in children and adolescents, which to our knowledge has not been reported in other studies with CoronaVac^®^ or mRNA vaccines against SARS-CoV-2. Compared to the pre-immune samples, we observed a significant increase in CD4^+^ T cell activation four weeks after the second dose of CoronaVac^®^ upon stimulation with four Mega-pools S, R, M, and N (**Figure 3A-B**). A significant increase in the activation of CD4^+^ T cells was found in 12-17 aged groups for all the MPs evaluated. In contrast, in the 3-11 years old group, a significant increase in the activation of CD4^+^ T cells with the stimulus S and N was found (**Figure 3C-F**). Additionally, the induction of memory cells induced by vaccination two weeks after the second dose compared to the pre-immune sample was analyzed (**Figure 4A-B**). An increase in the ratio of memory cells with respect to the pre-immune sample was observed in participants aged 12-17 years in the presence of all stimuli (**Figure 4C-F**). For the group 3-11 years, an increase in the ratio of memory cells was observed with respect to the pre-immune sample only in the presence of the S and N stimuli (**Figure 4F**). We see an increase in IFN-γ production by ELISPOT upon stimulation with S, R and N MPs after the second dose in the age group of 12-17 years old, but this is not significant as compared to the pre-immune sample (**Suppl. Fig 3A-D**). In addition, we did not observe any changes in the secretion of IL-4 using ELISPOT in response to SARS-CoV-2 MPs (**Suppl. Figure 3E-H**).

**Figure 3.**
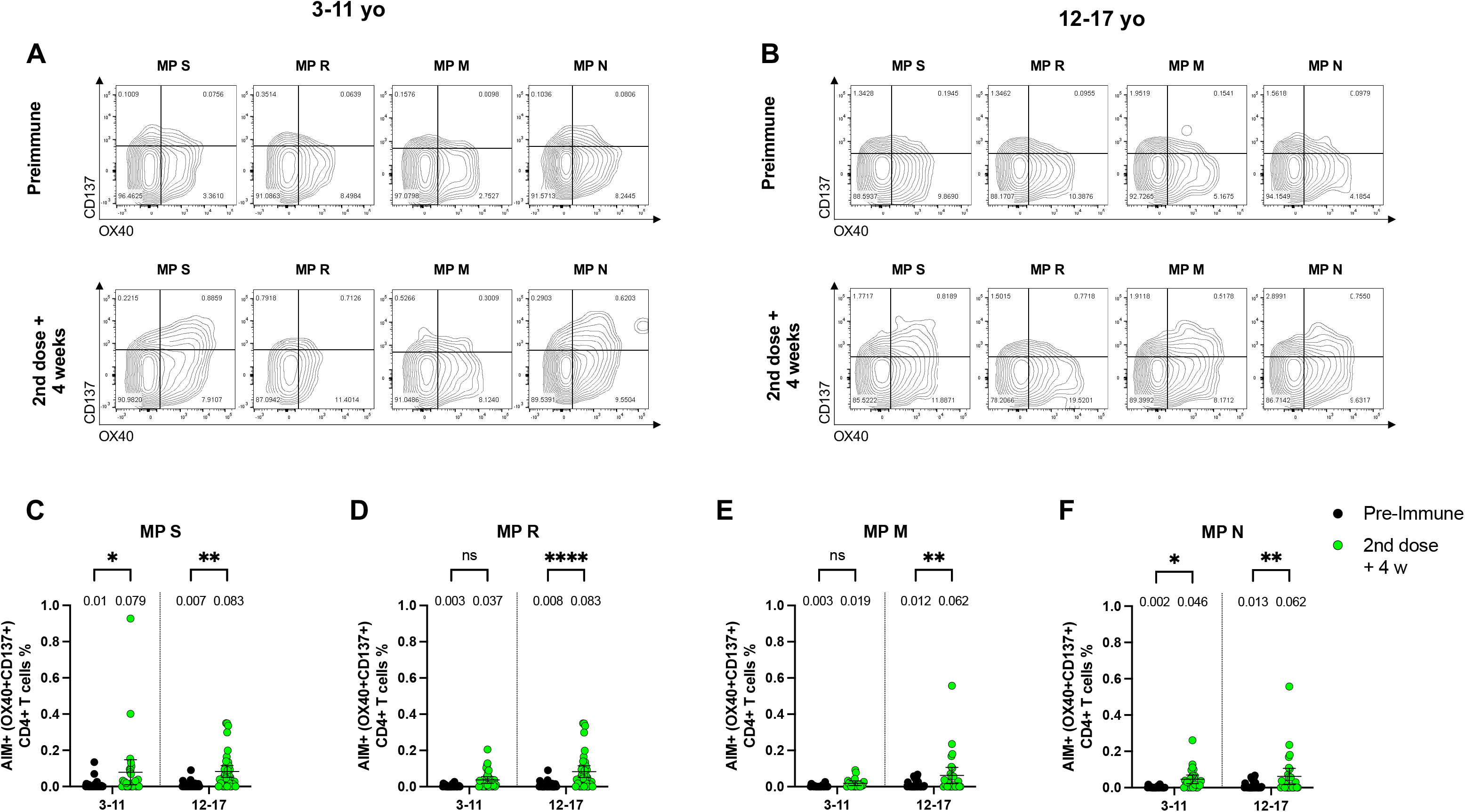
Changes in activation-induced markers (AIMs) expression in CD4^+^ T cells specific for SARS-CoV-2 after two doses of CoronaVac^®^ in children and adolescents. AIM^+^ CD4^+^ T cells were quantified in peripheral blood mononuclear cells of participants that received two doses of CoronaVac^®^ by flow cytometry, upon stimulation with mega-pools (MP) of peptides derived from SARS-CoV-2 proteins. The percentage of activated AIM^+^ CD4^+^ T cells (OX40^+^, CD137^+^) were determined upon stimulation for 24h with MPs S, R, M and N in pre-immune samples and samples obtained four weeks after the second dose. Data from flow cytometry was normalized against DMSO and analyzed separately by a Wilcoxon test against the pre-immune sample. Representative flow cytometry plots for participants aged 3-11 years old**(A)** and 12-17 years old **(B)** are shown. Percentage of AIM^+^ CD4^+^ T cells against the MPs S **(C)**, R **(D)**, M **(E)** and N **(F)** were obtained from a total of thirty participants aged 3-11 years old and thirty participants aged 12-17 years old. A two-way ANOVA was used to compare the percentage of AIM^+^ CD4^+^ T cells four weeks after the second dose against the pre-immune in both age groups. *p<0.5, **p<0.005, ****p<0.0001, n.s. non-significant.

**Figure 4.**
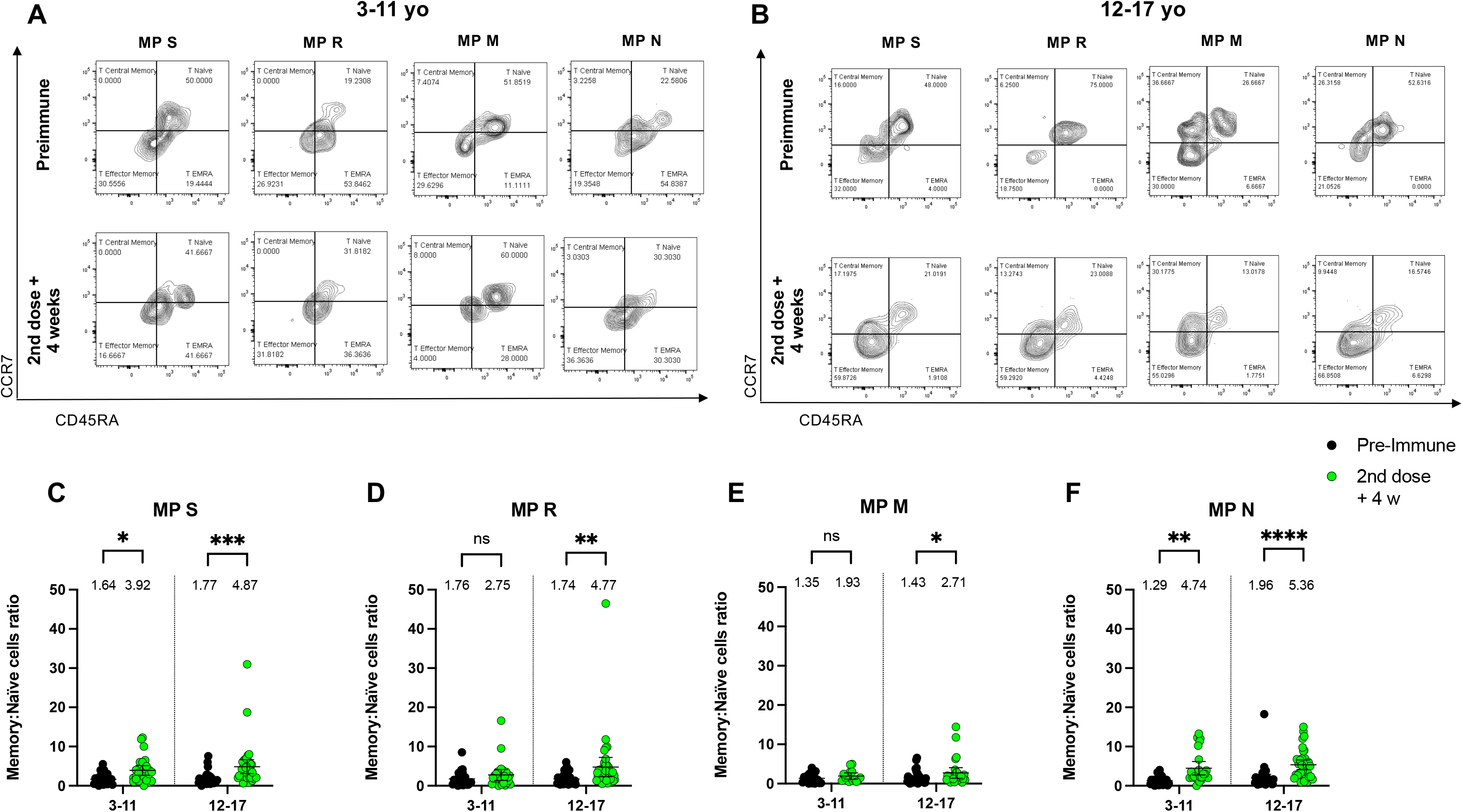
Changes in memory AIM^+^ CD4^+^ T cells specific for SARS-CoV-2 after two doses of CoronaVac^®^ in children and adolescents. Memory AIM^+^ CD4^+^ T cells were quantified in peripheral blood mononuclear cells of participants that received two doses of CoronaVac^®^, upon stimulation with mega-pools of peptides derived from SARS-CoV-2 proteins. The percentage of memory activated AIM^+^ CD4^+^ T cells (OX40^+^, CD137^+^, CD45RA^-^, CCR7^+/-^) were determined upon stimulation for 24h with MPs S, R, M and N in samples obtained at pre-immune and four weeks after the second dose. Data from flow cytometry was normalized against DMSO and analyzed separately by a Wilcoxon test against the pre-immune sample. Representative flow cytometry plots for participants aged 3-11**(A)** years old and 12-17 years old **(B)** are shown. Memory AIM^+^ CD4^+^ T cells against the MPs S **(C)**, R **(D)**, M **(E)** and N **(F)** were obtained from a total of thirty participants aged 3-11 years old and thirty participants aged 12-17 years old. A two-way ANOVA was used to compare the percentage of memory AIM^+^ CD4^+^ T cells four weeks after the second dose against the pre-immune sample in both age groups. *p<0.5, **p<0.005, ***p<0.001, ****p<0.0001, n.s. non-significant.

On the other hand, we did not observe an increase in CD8^+^ AIM^+^ T cells with MP CD8A and CD8B for participants aged 3 to 11 years or 12 to 17 years following the second dose of CoronaVac^®^, as compared to the pre-immune sample (**Data not shown**). Only a significant increase in memory CD8^+^ AIM^+^ T cells after vaccination with CoronaVac^®^ was only observed upon stimulation with MP CD8A in the age group of 3-11 years old (**Suppl. Figure 4C**).

### Two doses of CoronaVac® in children and adolescents promote the secretion of cytokines related to an antiviral profile

Secretion of the cytokines IL-2 and IFN-γ were evaluated using Luminex^®^ in PBMCs stimulated with MPs of peptides. We observed a significant increase in IL-2 secretion in response to the S and R MPs and M and N MPs for participants aged 12-17 years (**Figure 5A-D**). In the case of the 3-11 group, we observed a significant increase in response to the S, M, and N MP. In contrast, we did not observe a significant increase in IFN-γ release after the MP stimulation for participants aged 12-17 years (**Figure 5E-H**). However, participants between 3-11 years present a significant increase in the production of IFN-γ in response to the S, M, and N MPs (**Figure 5E and 5H**).

**Figure 5.**
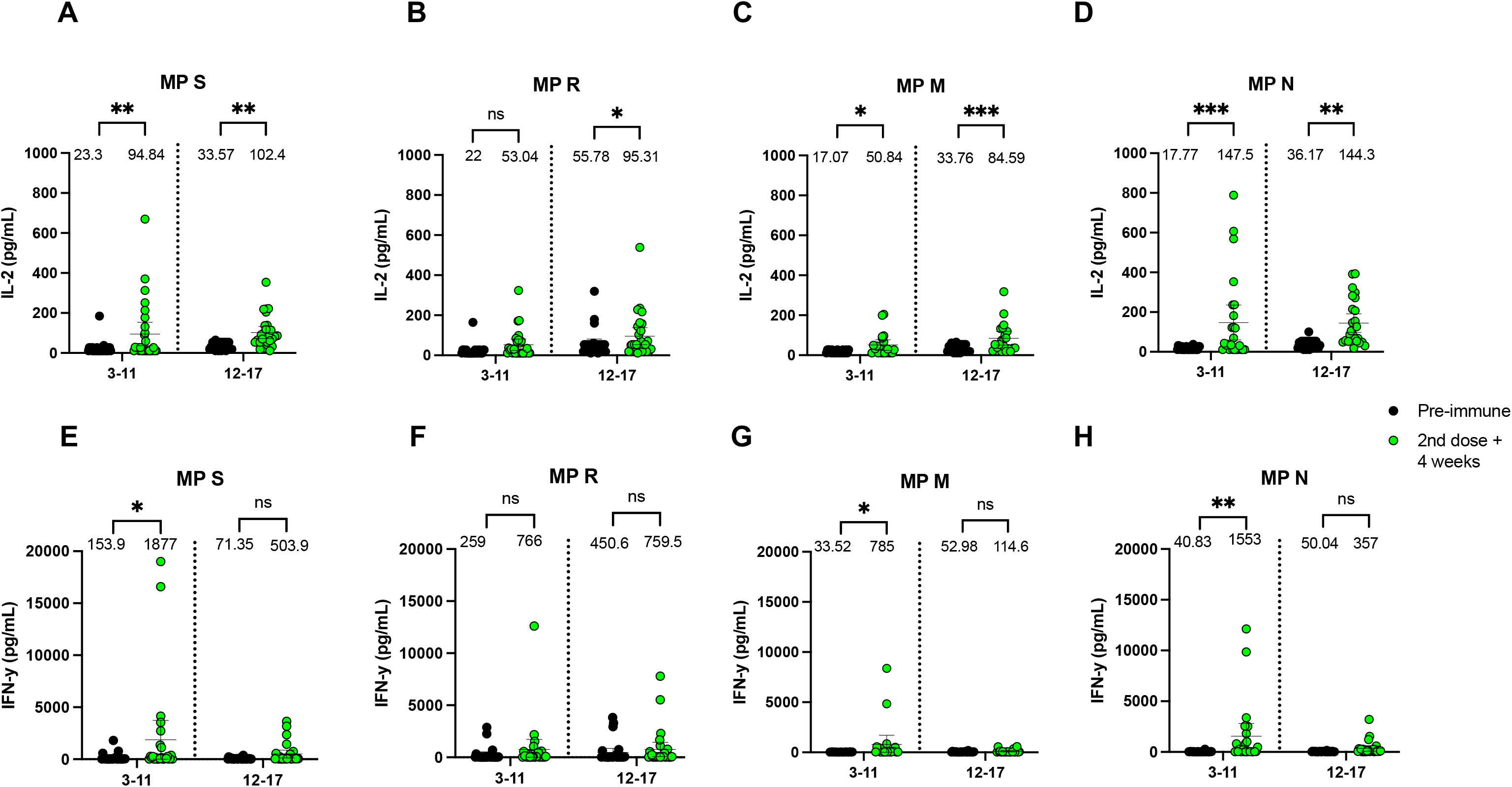
Changes in IL-2 and IFN-γ secretion by PBMCs stimulated with SARS-CoV-2 megapools of peptides after two doses of CoronaVac^®^ in children and adolescents. IL-2 and IFN-γ secretion was quantified in supernatants of peripheral blood mononuclear cells of participants that received two doses of CoronaVac^®^, upon stimulation with mega-pools of peptides derived from SARS-CoV-2 proteins for 18h by Luminex. Levels of IL-2 secretion against the MPs S **(A)**, R **(B)**, M **(C)** and N **(D)** and IFN-γ secretion against the MPs S **(E)**, R **(F)**, M **(G)** and N **(H)** are shown from a total of twenty-three participants aged 3-11 years old and twenty-three participants aged 12-17 years old. A two-way ANOVA was used to compare the level of cytokines four weeks after the second dose against the pre-immune sample. *p<0.5, **p<0.005, ***p<0.001, n.s. non-significant.

### Neutralizing antibodies and specific T cells induced by two doses of CoronaVac® in children and adolescents recognize Delta and Omicron variants of SARS-CoV-2

To assess whether CoronaVac^®^ induces immune responses against SARS-CoV-2 variants of concern, we evaluated by a pseudotype virus neutralization assay (pVNT) the neutralizing antibody production against variants of concern Delta and Omicron as compared to D614G (**Figure 6**). A 1.9-fold reduction relative to strain D614G (GMT 265.4, 95% CI=213.1-330.5) was found in neutralization against Delta (GMT 141.6, 95% CI=113.6-176.5), while 15.8 -fold reduction was identified against Omicron (GMT 16.8, 95% CI=13.95-20.26) (**Figure 6A**). The percentages of seropositivity show that an important reduction is observed for the Omicron variant (**Suppl. Table 3**). When we compare the response between both age groups, we do not find significant differences in any of the variants evaluated (**Figure 6B**). However, a significant mild reduction of AIM^+^ T cell against MP-S of the Delta variant (1.67-fold reduction) and a significant mild increase against MP-S of the Omicron variant (1.63-fold increase) was observed, as compared to the response obtained for MP-S of the wild type (WT) strain (**Figure 6C**). This T cell response was equivalent in both age groups (**Figure 6D**).

**Figure 6.**
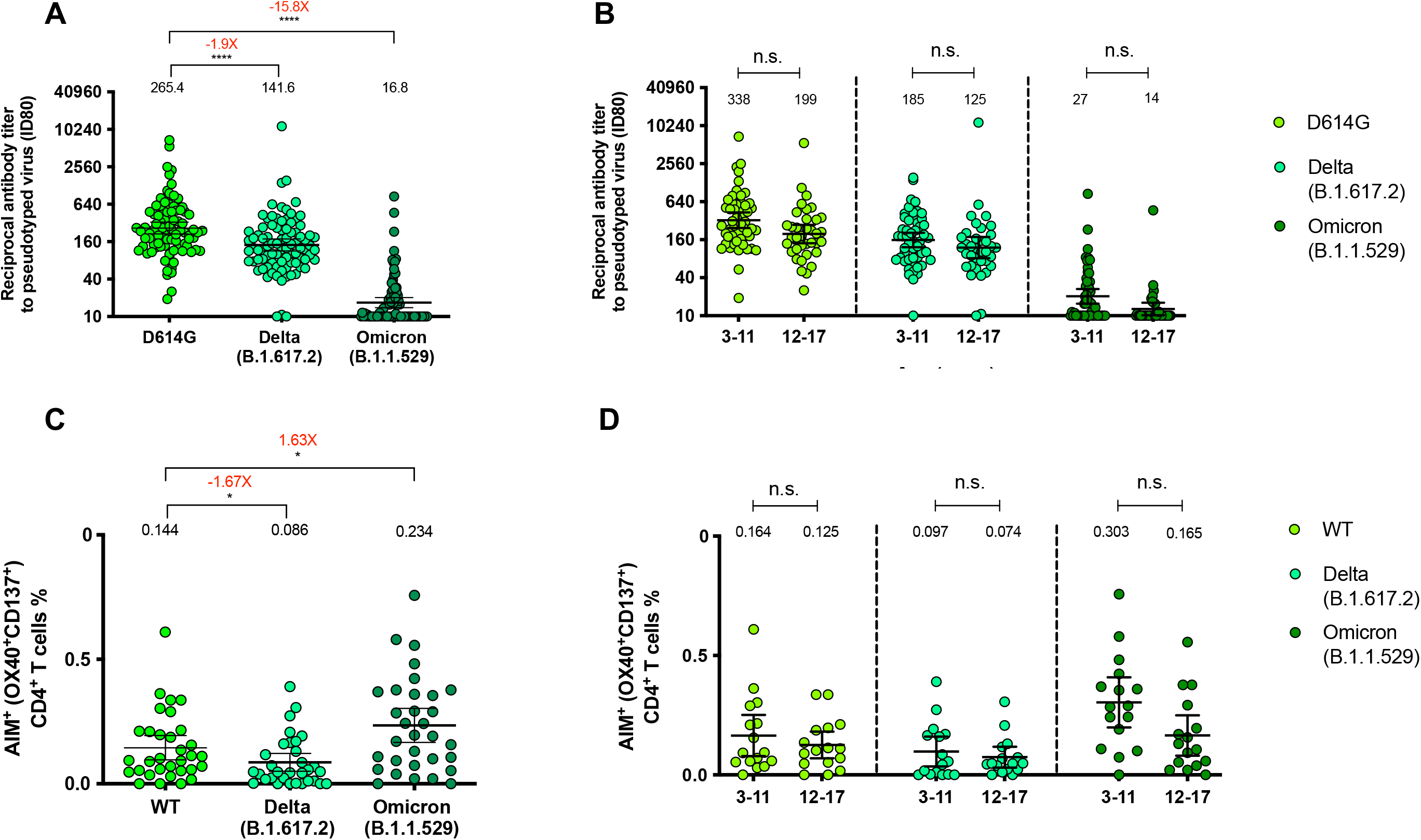
Quantification of circulating neutralizing antibodies and AIM^+^ CD4^+^ T cells against SARS-CoV-2 variants Delta and Omicron in participants that received two doses of CoronaVac®. (**A)** Neutralizing antibodies were detected in the plasma of eighty-eight participants, four weeks after the second dose of CoronaVac®, using a pseudotyped virus neutralization test (pVNT). Data are expressed as the reciprocal of the highest dilution preventing 80% of the infection (ID80). Numbers above the bars show the GMT, and the error bars indicate the 95% CI. The number above represents the fold change when it is compared with the response tof D614G variant. (**B)** Neutralizing antibody levels between fifty-two participants aged 3-11 and thirty-six participants aged 12-17 against D614G, Delta and Omicron variants are shown. (**C-D)** AIM^+^ CD4^+^ T cells against the variants Delta and Omicron were measured by flow cytometry. **(C)** Results were obtained from a total of thirty-two participants **(D)** Results are shown by age group (sixteen participants from each age group). A Friedman test assessed statistical differences of the mean to compare each variant against D614G whereas a two-way ANOVA was used to compare the two age groups for each variant. *p<0.05; ****p<0.0001, n.s. non-significant.

## Discussion

Previous studies have shown a 65.9% of effectiveness for immunization with two doses of CoronaVac^®^ in a 0-28 schedule ^23^. Here, we show that this vaccine has a very good safety profile, comparable to what was reported by Han^3^, being pain the main AE in both age groups but statistically higher in adolescents than in children. Most of the AEs were mild or moderate and no SAE related to the vaccine were reported.

Here we have also assessed the capacity of plasma samples from children and adolescents vaccinated with CoronaVac^®^ to neutralize SARS-CoV2, performing surrogate neutralizing antibody assays (sVNT), a pseudotyped virus (pVNT) assay, and conventional microneutralization assays in Vero E6 Cells (cVNT). Additionally, our study assessed T cell immunity ^3,4^ and immune responses against variants of concern Delta and Omicron. Two doses of CoronaVac^®^ administered in a 4-week interval stimulate the induction of both total and neutralizing antibodies in participants aged 3-17 years old four weeks after the second dose. This is the first report of total antibodies anti-S1-RBD expressed as WHO arbitrary units in children and adolescents vaccinated with CoronaVac^®^, allowing their comparison to other vaccine platforms, although clinical trials in children and adolescents have only reported neutralizing antibodies against SARS-CoV-2 ^4,5^. Compared to our previous work in an adult cohort vaccinated with two doses of CoronaVac^®^ in a 0-28 schedule, the total IgG measured as the geometric mean of arbitrary units of WHO (GMU) was 345.1 (95% CI=276.1-431.5) in adults aged ≥18 years old, four weeks after the second dose (unpublished results), whereas here we report GMU values of 964.9 (95% CI=503-1850) and 680 (95% CI=371-1245) in subjects aged 3-11 and 12-17 years old, respectively, suggesting an enhanced humoral response against SARS-CoV-2 in children and adolescents. In line with this, adults vaccinated with BNT162b2 and the mRNA-1273 exhibit GMU of 490.17 and 659, respectively, suggesting that children aged 3-11 vaccinated with CoronaVac^®^ exhibit higher levels of total anti-S1-RBD antibodies ^24^.

A phase 2 trial conducted in Chinese adolescents vaccinated with CoronaVac^®^ reported a GMT for neutralizing antibodies (with cVNT) of 146.0 four weeks after the second dose in 72 participants aged 12-17 years old, whereas here we report a GMT of 25.91 in 23 participants of the same age using cVNT. In the same study, children vaccinated with CoronaVac^®^ reported a GMT of 142.2 four weeks after the second dose in participants aged 3-17 years old, whereas here we report a GMT of 128.0 (95% CI=74.8-219.2) and 34.02 (95% CI=18.1-64.0) in 27 and 34 participants of 3-11 and 12-17 years, respectively (**Table 3**). The reported differences may be attributed to the use of different SARS-CoV-2 strains to perform such tests (D614G variant in the assays reported in this study). As a better comparison, clinical trials recently performed in healthy adults with neutralizing antibody titers reported as WHO arbitrary units (GMU) reported values of 178.2 (95% CI= 123.6-256.9) and 102.6 (95% CI=70.0-150.3) in participants aged 18-59 years old and older than 60 years old, respectively, for samples obtained four weeks after the second dose (also 0-28 schedule), which was measured by a sVNT assay^25^ . Here, using the same methodology, we found higher GMUs in adolescents aged 12-17 years old (492.2, 95% CI=342-708.3) and children aged 3-11 years old (713.1; 95% CI=565.8-898.8).

Our results suggest that CoronaVac^®^ promotes CD4^+^ T cell responses against SARS-CoV-2, which can be protective against infection and/or severe disease. Here we report a significant increase in CD4^+^ AIM^+^ T cells in response to S, R, M, and N MPs but no differences in CD8^+^ AIM^+^ T cells, in line with the results previously observed in adults vaccinated with two doses of CoronaVac^® 13^. We did not observe significant differences between age groups in CD4^+^ AIM^+^ T cells, suggesting that both children and adolescents can activate CD4^+^ T cell responses against SARS-CoV-2 following vaccination. Consistent with this, we show a significant increase in IL-2 secretion in response to S and N MPs in both age groups, whereas we detected a significant increase in response to the M MPs only in subjects aged 12-17 years old. Furthermore, we observed an increase in the frequency of memory CD4^+^ AIM^+^ T cells in response to SARS-CoV-2 MPs, although we observed a slightly higher induction of memory T cells in subjects aged 12-17 years old as compared to subjects aged 3-11 years old. These results agree with reports showing that memory T cell responses against SARS-CoV-2 structural proteins increase with age ^26^. As the formulation of CoronaVac^®^ contains the full-inactivated virus is important to understand whether the induction of cellular responses against viral antigens other than Spike may be important in conferring protection against severe disease. To our knowledge, this is the first study to report cellular immunity in children and adolescents vaccinated against SARS-CoV-2.

Moreover, we evaluated the neutralization using a pseudotyped virus using an ID80 against variants Delta and Omicron compared to the more ancestral strain D614G and found decreased antibody neutralization capacity against these variants. While we observed high seropositivity against D614G (100%), lower seropositivity against the variant Omicron was found (45.5%, Supp. Table 3) in line with previous reports indicating lower protection against variants of concern in adult cohorts after two doses of CoronaVac^® 13,19,27,28^. However, a booster dose of CoronaVac^®^ has been shown to increase virus neutralization of the variants of concern Gamma and Delta ^25^. Thus, it is possible that a booster dose of CoronaVac^®^ may be required to increase virus neutralization of circulating SARS-CoV-2 variants in children and adolescents, although this remains to be determined empirically. Previous studies performed in Israel showed a decrease in the transmission and the severe disease by SARS-CoV-2 twelve or more days after booster inoculation ^29^ and our previous study performed in adults showed that a booster dose of CoronaVac^®^ increases neutralization against SARS-CoV-2 WT strain and VOC Delta and Omicron ^25^.

On the other hand, we observed that both age groups elicited CD4^+^ AIM^+^ T cells in response to MPs from the variants Delta and Omicron. We observed a significant reduction in CD4^+^ AIM^+^ T cells against the Delta variant and, surprisingly, an increase against the Omicron variant. Several studies in vaccinated adults have shown that CD4^+^ T cell responses against variants of concern are conserved, and cross-reactive T cells against the Omicron variant have been reported ^22,31^. However, it is unclear why this pediatric population exhibit increased CD4^+^ AIM^+^ T cells against the Omicron variant, and further research is required to understand these results. Taken together, these results indicate that CoronaVac^®^ is safe in children and adolescents and induces both humoral and cellular responses able to recognize the variants of concern Delta and Omicron.

## Limitations

This study presents some limitations, such as samples obtained at few time points after vaccination, as compared to recent clinical trials performed in adults. In addition, infectious virus neutralization assays against the variants of concern Delta and Omicron needs to be performed to confirm the results obtained with the pVNT assay.

## Data Availability

All data produced in the present work are contained in the manuscript

## Acknowledgments

We would like to thank Rami Scharf, Jessica White, and Miren Iturriza-Gomara from PATH for their support on experimental design and discussion. We also thank the Vice Presidency of Research (VRI), the Direction of Technology Transfer and Development (DTD), the Legal Affairs Department (DAJ) of the Pontificia Universidad Católica de Chile. We are also grateful to the Administrative Directions of the School of Biological Sciences and the School of Medicine of the Pontificia Universidad Católica de Chile for their administrative support. We would like to thank the members of the independent data safety monitoring committee (members in the SA) for their oversight, and the subjects enrolled in the study for their participation and commitment to this trial.

Note: Members of the PedCoronaVac03CL Study Group and the Data and Safety Monitoring Board are listed in the Supplementary Appendix (SA).

---

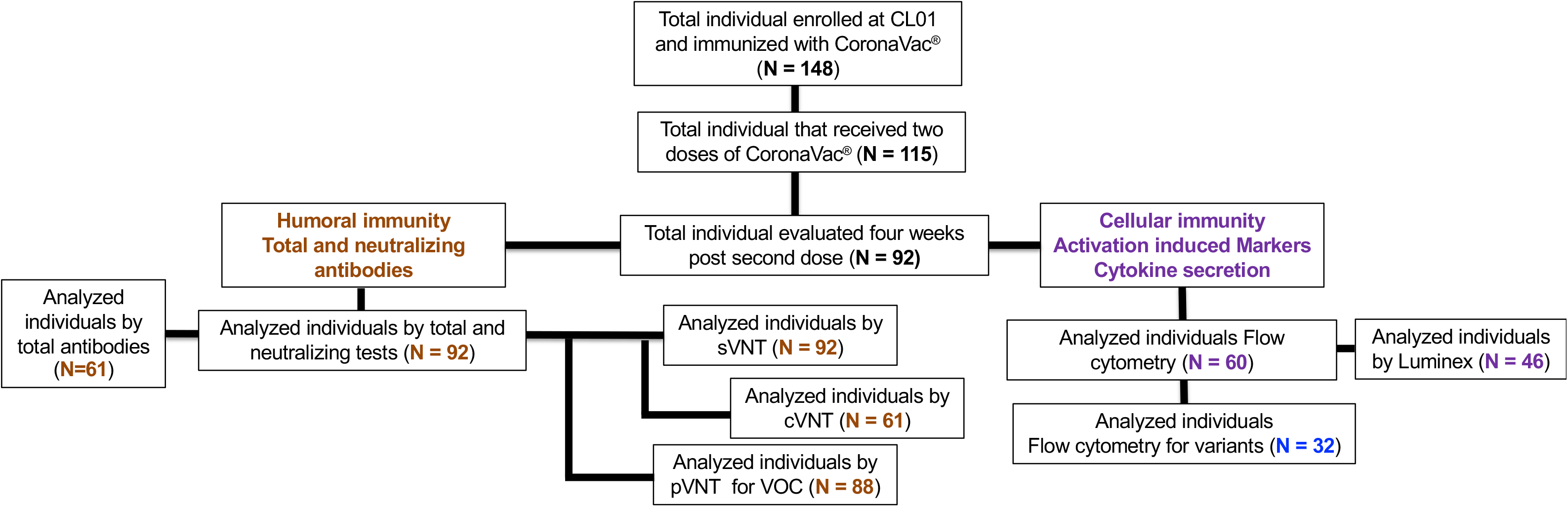

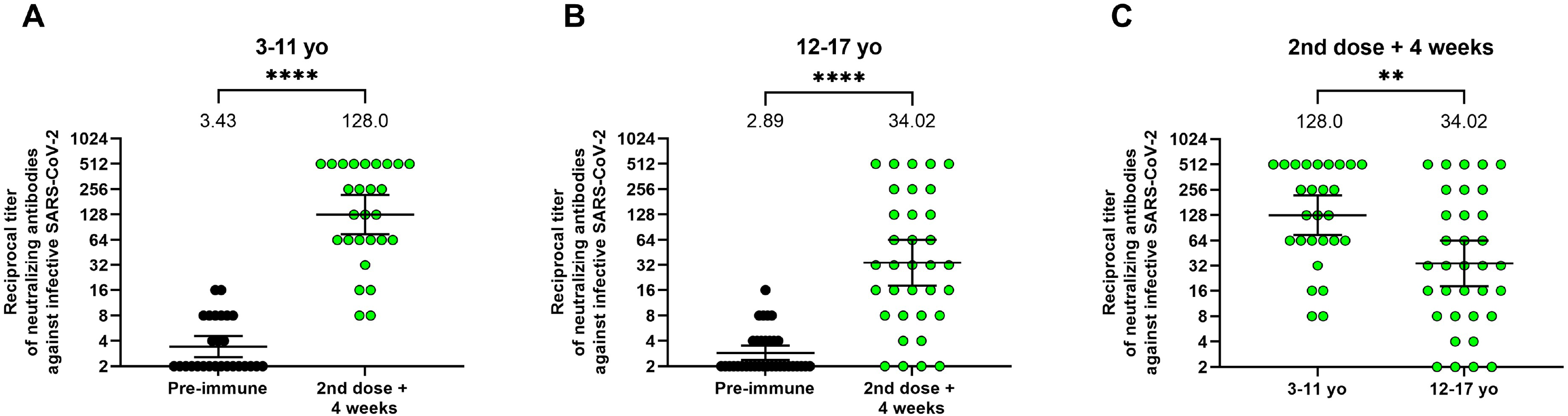

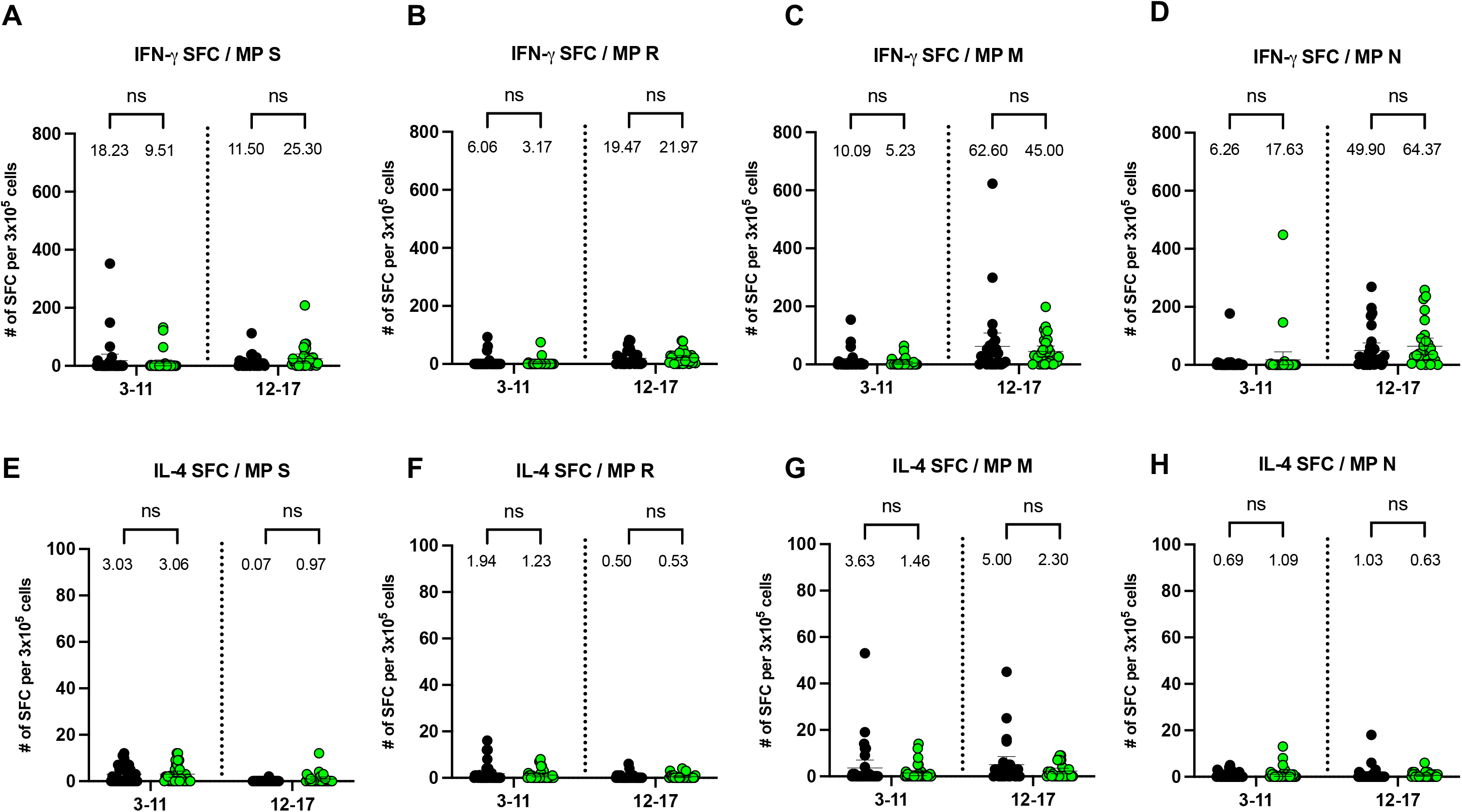

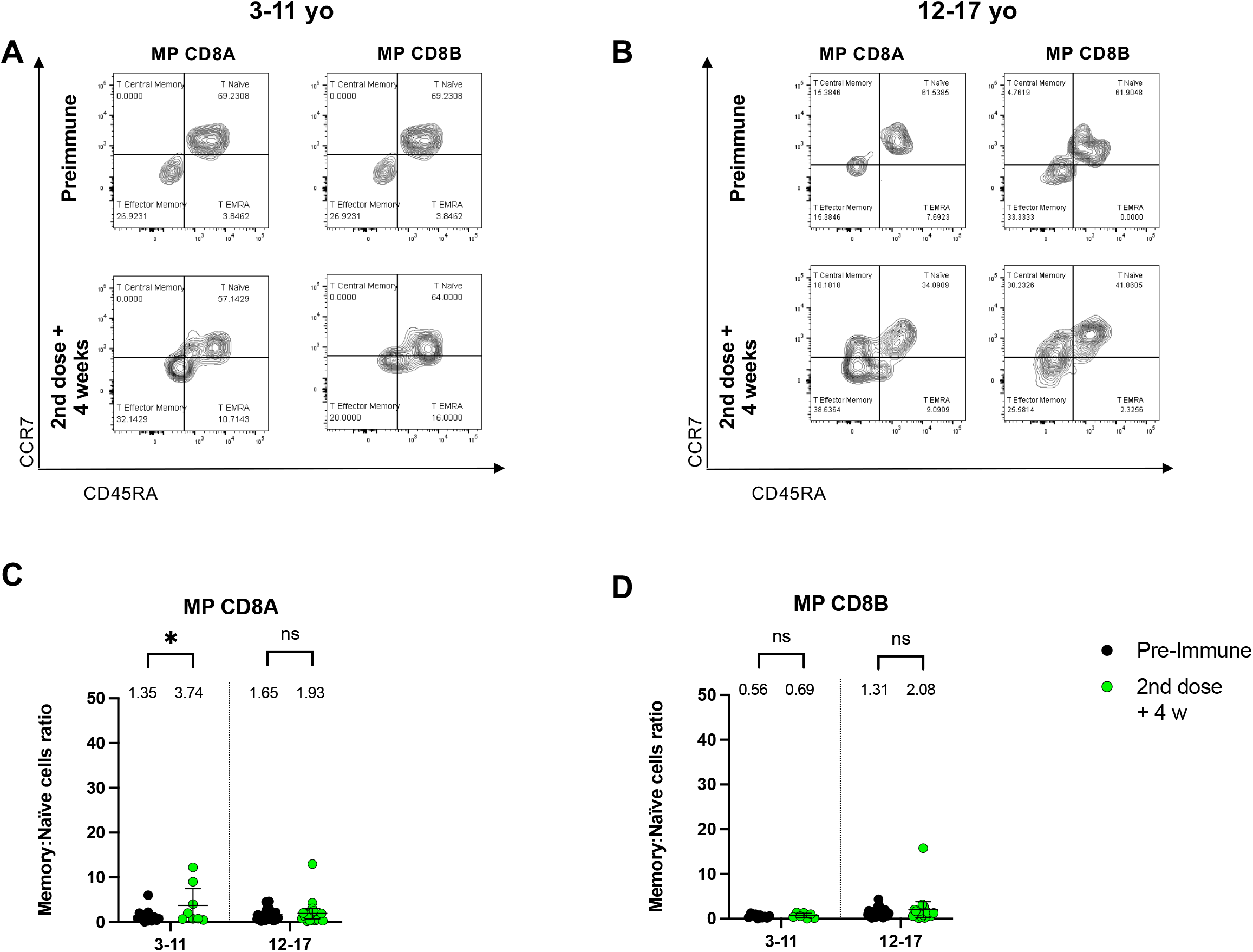

