## Supplementary material for "An inactivated SARS-CoV-2 vaccine is safe and induces humoral and cellular immunity against virus variants in healthy children and adolescents in Chile": Soto_et_al_Supplementary_Information_11Feb2022.docx

**Supplementary Information**

**Inclusion and Exclusion Criteria**

**Inclusion Criteria**

1.	Healthy children and adolescents aged 6 months to 17 years;

2.	The participants and/or their guardians are able to understand and sign the informed consent voluntarily (in accordance with the local regulations);

3.	Able to comply with study procedures based on the assessment of the Investigator;

4.	Female participants of childbearing potential (post-menarche girls or in accordance with the local standard of care) may be enrolled in the study if the participant fulfills all the following criteria:

•	Has a negative pregnancy test on the day of the first dose (Day 0).

•	Has practiced adequate contraception or has abstained from all activities that could result in pregnancy for at least 28 days prior to the first dose (Day 0).

•	Has agreed to continue adequate contraception through 3 months following the second dose (Day 28).

•	Is not currently breastfeeding.

5. Must be willing to provide verifiable identification (in accordance with the local regulations), has means to be contacted and to contact the investigator during the study.

**Exclusion Criteria**

Participants are excluded from the study if any of the following criteria apply:

- History of confirmed infection of SARS CoV-2 prior to randomization;
- History of contact with person infected with SARS-CoV-2 (has a positive nucleic acid test or an antigen test) within 14 days prior to randomization;
- Prior administration of an investigational or licensed coronavirus vaccine or current/planned simultaneous participation in another interventional study to prevent or treat COVID-19;
- Allergy to vaccines or vaccine/placebo ingredients, and serious adverse reactions to vaccines, such as urticaria, dyspnea, angioneuroedema;
- Personal or first-grade relative (siblings) history of multisystem inflammatory disease in children (MIS-C);
- Significant chronic illnesses that, in the opinion of the investigator, is at a stage where it might interfere with trial conduct or completion (may include, but are not limited to cardiovascular disease, liver or kidney disorders, respiratory illnesses)
- Significant chronic central nervous system diseases or neuromuscular disorders, psychosis or severe cognitive behavioral disorder, in the opinion of the investigator, including epilepsy, autism spectrum disorder, intellectual disabilities (excluding Down Syndrome);
- Acute central nervous system diseases such as encephalitis/myelitis, acute disseminating encephalomyelitis, and related disorders;
- History of autoimmune and/or haematological diseases (including but not limited to systemic lupus erythematosus, thyroidectomy, autoimmune thyroid disease, any form of malignant tumor, asplenia, functional asplenia, or splenectomy resulting from any condition); well controlled type I diabetes mellitus is allowed;
- History of bleeding disorders (e.g. factor deficiency, coagulopathy or platelet disorder), or prior history of significant bleeding or bruising following IM injections or venipuncture;
- Immunosuppressive therapy (systemic corticoid therapy, e.g. prednisone ≥2 mg/Kg/d or ≥20 mg/day for >14 days), cytotoxic therapy (antineoplastic chemotherapy, radiation therapy), (excluding topical or aerosol corticosteroid therapy) in the past 6 months;
- Receipt of blood products or immunoglobulins in the past 3 months;
- Receipt of other investigational drugs in the past 30 days;
- Receipt of attenuated live vaccines in the past 14 days;
- Receipt of inactivated or subunit vaccines in the past 7 days;
- Emerging of chronic diseases or acute exacerbation of stable chronic diseases (including but not limited to asthma, migraine, gastrointestinal disorder, etc.) prior to randomization;
- Acute febrile illness with oral temperature >37.6°C or axillary temperature >37.4°C on the day of vaccination (refer to section 7.1 Delay/Discontinuation of Study Vaccination); enrollment could be considered if the fever is absent for 72 hours;
- Any confirmed or suspected human immunodeficiency virus (HIV) infection;
- Children in care or under a court order;
- According to the investigator's judgment, the subject has any other factors that might interfere with the results of the clinical trial or pose additional risk to the subject due to participation in the study.

**Supplementary Tables**

**Supplementary Table 1. Frequency of local immediate adverse events by dose and age group.**

|  | 3-11 years (n=873) | 12-17 years (n=90) | 3-11 years (n=539) | 12-17 years (n=85) |
| --- | --- | --- | --- | --- |
|  | Dose 1 (n=963) | | Dose 2 (n=624) | |
|  | **Pain** | | | |
| 3-11 years | 33 (3.8) | | 9 (1.7) | |
| 12-17 years | 2 (2.2) | | 7 (8.2) | |
|  | **Redness** | | | |
| 3-11 years | 20 (2.3) | | 3 (0.6) | |
| 12-17 years | 0 | | 0 | |
|  | **Induration** | | | |
| 3-11 years | 5 (0.6) | | 0 | |
| 12-17 years | 1 (1.1) | | 0 | |
|  | **Pruritus** | | | |
| 3-11 years | 2 (0.2) | | 0 | |
| 12-17 years | 0 | | 1 (1.2) | |
|  | **Swelling** | | | |
| 3-11 years | 0 | | 0 | |
| 12-17 years | 1 (1.1) | | 0 | |
|  | **Other** | | | |
| 3-11 years | 1 (0.1) | | 1 (0.2) | |
| 12-17 years | 0 | | 0 | |

**Supplementary Table 2. Frequency of systemic immediate adverse events by dose and age group.**

|  | 3-11 years (n=873) | 12-17 years (n=90) | 3-11 years (n=539) | 12-17 years (n=85) |
| --- | --- | --- | --- | --- |
|  | Dose 1 (n=963) | | Dose 2 (n=624) | |
|  | **Headache** | | | |
| 3-11 years | 6 (0.7) | | 1 (0.2) | |
| 12-17 years | 2 (2.2) | | 1 (1.2) | |
|  | **Fatigue** | | | |
| 3-11 years | 4 (0.5) | | 1 (0.2) | |
| 12-17 years | 0 | | 0 | |
|  | **Muscle pain** | | | |
| 3-11 years | 1 (0.1) | | 0 | |
| 12-17 years | 0 | | 1 (1.2) | |
|  | **Nausea** | | | |
| 3-11 years | 1 (0.1) | | 0 | |
| 12-17 years | 0 | | 0 | |
|  | **Skin or mucosa abnormality** | | | |
| 3-11 years | 0 | | 0 | |
| 12-17 years | 0 | | 1 (1.2) | |

| **Test** | **Parameter** | **Variant** | | |
| --- | --- | --- | --- | --- |
|  |  | **D614G** | **Delta** | **Omicron** |
| Neutralizing antibodies (pVNT) | Seropositivity n/N | 88/88 | 86/88 | 40/88 |
|  | (%) | 100 | 97.7 | 45.5 |
|  | GMT | 265.4 | 141.6 | 16.81 |
|  | 95% CI | 213.1-330.5 | 113.6-176.5 | 14.0-20.3 |

**Supplementary Table 3: Seropositivity rates, Geometric Mean titers (GMT) of circulating antibodies against variant of concern of SARS-CoV-2.**

**Supplementary Figure Legends**

**Suppl. Figure 1. Sample size included in immunogenicity assays.** From the total number of participants enrolled in one clinical center (CL01, Marcoleta), one hundred and fifteen received two doses of CoronaVac^®^ in a four-week interval (0-28 days schedule of vaccination). Ninety-two of them were included in this study. Sixty-one were selected to analyze total antibodies by chemoelectroluminescence; ninety-two participants were tested for neutralizing antibodies by sVNT, sixty-one were tested for neutralizing antibodies by cVNT, and eighty-eight were selected to analyze neutralizing antibodies by pVNT. Sixty participants were selected to analyze cellular immunity by flow cytometry, thirty-two were analyzed for cellular responses against variants of concern by flow cytometry and forty-six were analyzed by Luminex.

**Suppl. Figure 2. CoronaVac^®^ immunization induces neutralizing antibodies in children and adolescents after two vaccine doses.** Neutralizing antibodies titers in plasma were evaluated using conventional virus neutralizing test (cVNT). The cytopathic effect was evaluated in twenty-seven children aged 3-11 years old (**A**) and thirty four adolescents aged 12-17 years old (**B**). Comparison between the titers of neutralizing antibodies in children and adolescents (**C**). Data is represented as Geometric Mean Titer (GMT) and the error bars indicate the 95% CI. A Wilcoxon test analyzed data to compare the levels of antibodies four weeks after the second dose against the pre-immune samples. ****p<0.0001.

**Suppl Figure 3. Expression levels of IL-4 and IFN-γ from specific T cells against MPs of SARS-CoV-2 after two doses of CoronaVac^®^ in children and adolescents**. IL-4 and IFN-γ secretion from peripheral blood mononuclear cells of volunteers that received two doses of CoronaVac^®^, upon stimulation with mega-pools of peptides derived from SARS-CoV-2 proteins for 48h by ELISPOT. Spot Forming Cells (SFC) from IFN-γ secretion against the MPs S **(A)**, R **(B)**, M **(C)** and N **(D)** and IL-4 secretion against the MPs S **(E)**, R **(F)**, M **(G)** and N **(H)** are shown from a total of twenty-three volunteers aged 3-11 years old and twenty-three volunteers aged 12-17 years old. A two-way ANOVA was used to compare used to compare the level of cytokines four weeks after the second dose against the pre-immune sample. ns: no significant.

**Suppl Figure 4. Changes in memory AIM^+^ CD8^+^ T cells specific for SARS-CoV-2 after two doses of CoronaVac**^®^ **in children and adolescents**. Memory AIM^+^ CD8^+^ T cells were quantified in peripheral blood mononuclear cells of volunteers that received two doses of CoronaVac^®^, upon stimulation with mega-pools of peptides derived from SARS-CoV-2 proteins. The percentage of memory activated AIM^+^ CD8^+^ T cells (CD69^+^, CD137^+^, CD45RA^-^, CCR7^+/-^) were determined upon stimulation for 24h with MPs S, R, M and N in samples obtained at pre-immune and four weeks after the second dose. Data from flow cytometry was normalized against DMSO and analyzed separately by a Wilcoxon test against the pre-immune. Representative flow cytometry plots for volunteers aged **(A)** 3-11 years old and **(B)** 12-17 years old are shown. Memory AIM^+^ CD8^+^ T cells against the MPs CD8A **(C)**, CD8B **(D)** were obtained from a total of thirty volunteers aged 3-11 years old and thirty volunteers aged 12-17 years old. A two-way ANOVA was used to compare the percentage of memory AIM^+^ CD8^+^ T cells four weeks after the second dose against the pre-immune sample in both age groups. *p<0.5, **p<0.005, ***p<0.001, ****p<0.0001.
