## Appendix for "An inactivated SARS-CoV-2 vaccine is safe and induces humoral and cellular immunity against virus variants in healthy children and adolescents in Chile": Soto_et_al_Supplementary_Appendix_15Feb2022.docx

**1. Multicenter Study Group**

1. Patricio Astudillo Paredes
2. Epifanía Hernández Jara
3. Héctor Morán Fernández
4. Javiera Arenas Urra
5. Stephani Ascui Baeza
6. María Olivia Cabrera
7. José Romero Muñoz
8. Gonzalo Alarcón Andrade
9. Rocío Rodríguez Espósito
10. Anwar Gutiérrez Silva
11. Fernanda Pérez Gutiérrez
12. Alma Muñoz Muñoz

**Center CL02: San Carlos de Apoquindo – Pontificia Universidad Católica de Chile**

1. Marcela Potin Santander
2. Sofia López
3. Tania Weil

**Center CL03: Centro medico San Joaquín - Pontificia Universidad Católica de Chile**

1. Macarena Goldsack
2. Deidyland Arenas
3. Andrea Araya
4. Javiera Moore
5. Lorena Pilicita
6. Vania Valenzuela
7. Catalina Campos
8. Mauricio Soto

**Center CL05: Clínica Alemana – Universidad del Desarrollo**

1. Andrea Schilling
2. Alberto Trautmann,
3. Ana Fritis,
4. Daniela Pavez,
5. Javiera Arancibia,
6. Lilian Rubio,
7. Paula Viviani
8. Vinka Basic.
9. Cassandra Cárcamo

**Center CL06: Clínica Valdivia - Universidad Austral**

1. Mario Calvo Gil
2. Marisol Wenzel
3. Nicole Carey
4. Roberto Burgos.

**Center CL07: Hospital de Puerto Montt**

1. Loreto Twele
2. Daniel Beltrán
3. Silvana Grandón
4. Carlos Tovar

**Center CL08: Hospital Gustavo Fricke – Universidad de Valparaíso**

1. Marcela González
2. Daniela Fuentes
3. Teresa Ramírez
4. Mariela Cepeda Corrales
5. Nataly Martínez López

**Center CL09: Complejo Asistencial Dr. Sotero del Rio**

1. Valentina Gutiérrez
2. Felipe Reyes
3. Armando Lavayen
4. Melissa González
5. Monserrat Gutiérrez
6. Noris Rengifo
7. Carla Ortega
8. Florencia Saver

**Center CL10: Hospital Roberto de Río – Universidad de Chile**

1. Lorena Tapia
2. Mirta Acuña
3. Javiera Albornoz
4. Tania Cariqueo
5. Alejandro Velásquez
6. Yennybeth Leiva Chamorro

**Center CL11: Hospital Exequiel Gonzalez Cortés - U de Chile**

1. Rodolfo Villena.

**Center CL12: Hospital Clínico Universidad de Antofagasta – Universidad de Antofagasta**

1. Antonio Cárdenas
2. Angello Retamal
3. Carmen Ludeña
4. Carolina Hermosilla
5. Gustavo Keilhold
6. Francisco Cammarata-Scalisi
7. Jessica Álvarez
8. Jessica Romero
9. Pía Villarroel
10. Francisca Muñoz
11. Jorge Maya
12. Andrés Canales
13. Margarita K. Lay
14. Christian Muñoz
15. Roylester Araya

**2. Members of the Independent Data Safety Monitoring Committee.**

- Xuanyi Wang, Research Scientist, Institutes of Biomedical Sciences, Fudan University, Shanghai, China
- Lorenz von Seidlein, Asc. Professor, Mahidol-Oxford Tropical Medicine Research Unit, Bangkok, Thailand
- Xia Jielai, professor and doctoral supervisor of Health Statistics, Department of military preventive medicine, Air Force Medical University
- Dongliang Yang, MD. Director of Sino-German "International Joint Laboratory of Viral Infection and Immunity", the Director of International Cooperative Base for Infection and Immunity of Hubei Province, China
- Carlos M. Perez, MD. Dean and Professor of Medicine, Faculty of Medicine and Science. Universidad San Sebastián, Chile.
- Zhaolong Cao, M.D.Ph.D. Head Physician, Professor of pulmonary and critical medicine at university of Wuhan, medical college
